# Identifying Sequential Complication and Mortality Patterns in Diabetes Mellitus: Comparisons of Machine Learning Methodologies

**DOI:** 10.1101/2020.12.21.20248646

**Authors:** Jiandong Zhou, Sharen Lee, Wing Tak Wong, Tong Liu, Leonardo Roever, Kamalan Jeevaratnam, William KK Wu, Ian Chi Kei Wong, Gary Tse, Qingpeng Zhang

## Abstract

**Background:** Diabetes mellitus-related complications adversely affect the quality of life. Better risk-stratified care through mining of sequential complication patterns is needed to enable early detection and prevention.

**Methods:** Univariable and multivariate logistic regression was used to identify significant variables that can predict mortality. A sequence analysis method termed Prefixspan was applied to identify the most common couple, triple, quadruple, quintuple and sextuple sequential complication patterns in the directed comorbidity pathology network. A knowledge enhanced CPT+ (KCPT+) sequence prediction model is developed to predict the next possible outcome along the progression trajectories of diabetes-related complications.

**Findings:** A total of 14,144 diabetic patients (51% males) were included. Acute myocardial infarction (AMI) without known ischaemic heart disease (IHD) (odds ratio [OR]: 2.8, 95% CI: [2.3, 3.4]), peripheral vascular disease (OR: 2.3, 95% CI: [1.9, 2.8]), dementia (OR: 2.1, 95% CI: [1.8, 2.4]), and IHD with AMI (OR: 2.4, 95% CI: [2.1, 2.6]) are the most important multivariate predictors of mortality. KCPT+ shows high accuracy in predicting mortality (F1 score 0.90, ACU 0.88), osteoporosis (F1 score 0.86, AUC 0.82), ophthalmological complications (F1 score 0.82, AUC 0.82), IHD with AMI (F1 score 0.81, AUC 0.85) and neurological complications (F1 score 0.81, AUC 0.83) with a particular prior complication sequence.

**Interpretation:** Sequence analysis identifies the most common pattern characteristics of disease-related complications efficiently. The proposed sequence prediction model is accurate and enables clinicians to diagnose the next complication earlier, provide better risk-stratified care, and devise efficient treatment strategies for diabetes mellitus patients.

## Introduction

Diabetes mellitus is a global problem and its associated expenditure is forecasted to rise [1]. Moreover, disease-related complications adversely affect the quality-of-life and treatment for diabetic patients with complications are costlier than for those without complications [2, 3]. Therefore, there is a need for better risk-stratified care, which would enable early complication detection and prevention [4, 5]. Most diabetes-related complications take several years to develop [6]. To aid clinical decision making, it is important to identify the typical trajectories of disease progression [7]. This would allow clinicians to identify complications early and devise effective treatment strategies.

However, most studies in diabetes are often limited to disease onset prediction without consideration of their temporal patterns. Whilst a 2017 Korean study from have applied network analysis to illustrate associations between diagnostic disease pairs [8], studies on temporal trajectory clusters of diseases are limited to two studies only, from the Korea [9] and Denmark [10]. Moreover, there has been no study to date on the trajectory patterns of diabetic-related complications specifically. In this study, therefore, we examine the trajectory patterns of diabetic complications using a technique called knowledge enhanced compact prediction tree plus (KCPT+) in diabetic patients who are on insulin therapy.

## Methods

### Study population and definitions

The study was approved by The Joint Chinese University of Hong Kong – New Territories East Cluster Clinical Research Ethics Committee and Institutional Review Board of the University of Hong Kong/Hospital Authority Hong Kong West Cluster. This was a territory-wide retrospective observational cohort study of diabetes mellitus patients who presented to outpatient clinics of the Hospital Authority of Hong Kong and are prescribed insulin, between 1^st^ January and 31^st^ December 2009. Through the Clinical Data Analysis and Reporting System (CDARS), a healthcare database that integrates patient information across all 43 publicly funded hospitals and their associated ambulatory and primary care facilities in Hong Kong to establish comprehensive medical records. The system has been used by multiple research teams, including our team, for epidemiological research in the past [11-14].

### Data of individual patient data and outcomes

Baseline characteristics of the patients were obtained from CDARS: 1) age, 2) gender, 3) diabetes type, 4) pre-existing comorbidities of chronic renal disease (CKD), chronic obstructive pulmonary disease (COPD), chronic liver disease (CLD), heart failure (HF), ischemic heart disease (IHD), hypertension, acute myocardial infarction (AMI) and stroke. Details on cardiovascular and anti-diabetic medications were also extracted.

Clinical outcomes, patient characteristics, and pharmacological treatment details were extracted. The patient outcomes from January 1^st^, 2009 to December 31^st^, 2013 were extracted. The primary outcome is all-cause mortality, and the secondary outcomes, as defined by their International Classification of Disease, Ninth Edition (ICD-9) codes (**Supplementary Table 1**), include: 1) neurological, ophthalmological and renal diabetic complications, 2) dementia, 3) osteoporosis, 4) peripheral vascular disease (PVD), 5) intracranial hemorrhage, 6) ischemic stroke and transient ischemic attack (TIA), 7) IHD with AMI, IHD without AMI, AMI without known IHD, and heart failure (HF), 8) atrial fibrillation (AF) (**Table 1**).

**Table 1.** Age characteristics of patients with diabetic complications.

|  | All patients<br>Median [IQR] | Male patients<br>Median [IQR] | Female patients<br>Median [IQR] | P value |
| --- | --- | --- | --- | --- |
| Ophthalmological | 65.9(58.0-75.0);n=4705 | 64.4(57.0-73.0);n=2385 | 67.5(59.0-76.0);n=2320 | <0.0001*** |
| Neurological | 67.8(60.0-76.0);n=1861 | 65.6(58.0-74.0);n=1110 | 71.3(62.0-79.0);n=751 | <0.0001*** |
| Renal | 71.0(61.0-78.0);n=5389 | 69.0(59.0-77.0);n=2853 | 73.1(64.0-80.0);n=2536 | <0.0001*** |
| Peripheral vascular disease | 71.7(62.0-80.0);n=712 | 69.2(59.0-78.0);n=412 | 76.1(67.0-81.0);n=300 | <0.0001*** |
| IHD without AMI | 71.9(63.0-79.0);n=2869 | 69.7(61.0-78.0);n=1529 | 74.1(66.0-81.0);n=1340 | <0.0001*** |
| IHD with AMI | 73.2(64.0-81.0);n=2458 | 70.7(61.0-79.0);n=1237 | 75.8(67.0-82.0);n=1221 | <0.0001*** |
| Ischemic stroke | 75.1(66.0-82.0);n=1350 | 73.0(64.0-80.0);n=680 | 77.1(69.0-83.0);n=670 | <0.0001*** |
| Heart failure | 75.2(66.0-81.0);n=1810 | 73.5(65.0-81.0);n=1017 | 77.1(68.0-82.0);n=793 | <0.0001*** |
| Atrial fibrillation | 76.5(69.0-82.0);n=1846 | 74.8(67.0-81.0);n=876 | 77.8(71.0-83.0);n=970 | <0.0001*** |
| AMI without known IHD | 77.1(70.0-82.0);n=720 | 75.1(69.0-82.0);n=352 | 78.6(72.0-83.0);n=368 | 0.0002*** |
| Mortality | 77.3(69.0-84.0);n=8460 | 75.2(66.0-82.0);n=4415 | 79.2(72.0-85.0);n=4045 | <0.0001*** |
| Osteoporosis | 78.7(70.0-84.0);n=275 | 77.9(68.0-84.0);n=56 | 79.1(72.0-84.0);n=219 | 0.4350 |
| Dementia | 79.9(74.0-84.0);n=952 | 78.4(72.0-83.0);n=403 | 80.9(75.0-85.0);n=549 | <0.0001*** |

### Statistical analysis

Continuous variables were presented as median (95% confidence interval [CI] or interquartile range [IQR]) and categorical variables were presented count (%). The χ2 test with Yates’ correction was used for 2×2 contingency data, and Pearson’s χ2 test was used for contingency data for variables with more than two categories. The Mann-Whitney U test was used to compare continuous variables. Differences between groups were tested using Kruskal-Wallis analysis of one-way variance (ANOVA). For each category of complication, we compared the age of onset and the difference between male and female groups. A two-sided α of less than 0.05 was considered statistically significant. Prefixspan [15] was used to extract the sequential patterns of complications. To identify the significant complication factors associated with mortality of these diabetes mellitus patients, univariate logistic regression was used to estimate odds ratios (ORs) and 95% CIs. To avoid overfitting in the model, significant univariable variables previously identified were chosen for multivariable analysis. Statistical analyses were performed using RStudio software (Version: 1.1.456) and Python (Version: 3.6). Experiments are simulated on a 15-inch MacBook Pro with 2.2 GHz Intel Core i7 Processor and 16 GB RAM.

### Network analysis of comorbidities

The following properties of the comorbidity network were extracted: connection degree (including in and out degree measures), eccentricity [16], closeness centrality [17], harmonic closeness centrality [18], betweenness centrality [19], eigen centrality [20], hub [21], PageRank [22], and clustering coefficient [23]. Centrality measures identify the most important nodes in a network. Closeness centrality indicates how close a node is to all other nodes in the network, calculated as the average of the shortest path length from the node to every other node in the network. Harmonic closeness centrality (also known as valued centrality) which is a variant of closeness centrality. Betweenness centrality captures how much a given node is in-between others and is measured with the number of shortest paths (between any couple of nodes in the graphs) that passes through the target node. Betweenness measure is moderated by the total number of shortest paths existing between any couple of nodes of the network. Eccentricity centrality is a measure of the centrality of a node in a network based on having a small maximum distance from a node to every other reachable node (i.e. the graph eccentricities). The measures of hub are also used to indicate node importance in the network. PageRank measures the transitive influence or connectivity of nodes, and its main difference from eigen centrality is that it accounts for link direction. The concept of the eigenvector centrality of a node is that the centrality index is determined not only by its position in the network but also by the neighboring nodes. Clustering coefficient is a measure of the degree to which nodes in a graph tend to cluster together. We interpreted the network properties of the complication outcomes in order to detect their roles in the sequential pathology network.

### Development of an accurate sequence prediction model

One of the important and meaningful tasks for complication sequence analysis in diabetes mellitus is to predict the next possible complication outcome of a patient based on his/her previous complications. In this study, we developed an accurate sequence prediction model that can accurately predict the next complication outcome (or mortality) of a (sub)sequence based on the previously observed complications. For instance, if a patient had sequential complications of renal complication, heart failure, and ischemic stroke at age 68, 75, and 76, respectively, it would be important to predict the next most likely complication (or mortality) to allow for early detection and personalized treatment strategy. The problem is thus a typical supervised many-to-one sequence prediction problem.

### Sequential pattern analysis

Compact prediction tree plus (CPT+) has been proposed as a fairly new probabilistic predictive model to assist sequential pattern analysis [24, 25]. In this study, we developed a knowledge enhanced CPT+ (KCPT+) model which further improves overall prediction ability by considering previously known onset probability of couple, triple, and quadruple sequences, and at the same time remain the advantage of CPT+ to capture the subsequence similarities without information loss.

Specifically, for modeling contribution, we first conduct preliminary sequence analysis and identified the onset probabilities of couple, triple, and quadruple complication sequences in the diabetes mellitus dataset, which provides a broad prior understanding of more frequently occurred complication sequences. Then we incorporated these important prior sequence onset estimations into the optimization process of CPT+ model, to increase the probability of generating the next complication outcome if it is contained in a sequence (couple, triplet, or quadruple, quintuple and sextuple) that has been known to happen more frequently. In contrast, the predicted probability of a complication outcome is decreased if it is in a sequence that has a low onset possibility.

Most patients with diabetes mellitus had multiple complications throughout their lifetime. The model training and testing consider mortality and other complication as primary outcomes to be predicted based on the input of former complication sequences before the onset of the outcome. For instance, the model can be used to distinguish patients that may suffer from the most severe outcome (i.e., mortality) and requires immediate medical assistance. The model can also be applied to other complication outcome predictions with the input of previous complication sequences. In this way, the model can predict the next outcome based on any given previously experienced complication of a patient with diabetes mellitus.

### Performance evaluation

To evaluate the model’s performance of predicting the outcomes of sequences, we use evaluation metrics of accuracy (ratio of true predictions over all sequence predictions), the precision, sensitivity/recall, F1 scores (defined below), Matthew’s correlation coefficient (MCC) and area under the curve (AUC) of the receiving operating characteristics (ROC) curve.

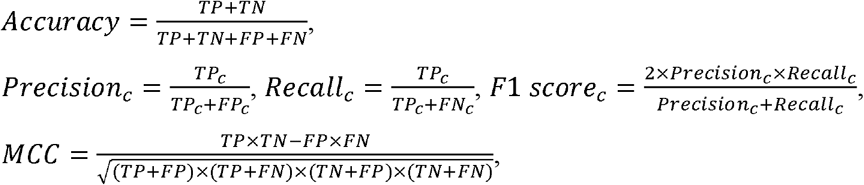

where *c* ∈ *C* represents the positive or negative class, *N* is the number of all sequences (couple, triplet, quadrable, quintuple and sextuple), *TN*_*c*_ in class *c, TP*_*c*_, *FP*_*c*_, and *FN*_*c*_ represent true positive, true negative, false positive, and false negative rates for class *c*, respectively. We compare the sequence prediction performance of the proposed KCPT+ model with baselines including CPT, recurrent neural network (RNN) [26], and long short-term memory network (LSTM) [27].

## Results

### Cohort characteristics

This study included a total of 14,144 diabetic patients (51% males). The descriptive statistics of complication onsets at different age intervals, stratified by gender, are shown in **Figure 1**. The median age of onset for different complications ranked in ascending order is shown in **Table 1**. Ophthalmological complication occurs the earliest with a median age of onset of 65.9 years old [58.0-75.0] (male: 64.4[57.0-73.0] and female: 67.5[59.0-76.0]), followed by neurological complications (median age 67.8 [60.0-76.0]; male: 65.6[58.0-74.0] and female: 71.3[62.0-79.0]), and renal complications (median age 71.0 [61.0-78.0]; male: 69.0[59.0-77.0] and female: 73.1[64.0-80.0]). The onset age for the remaining complications is detailed in **Table 1**.

**Figure 1.**
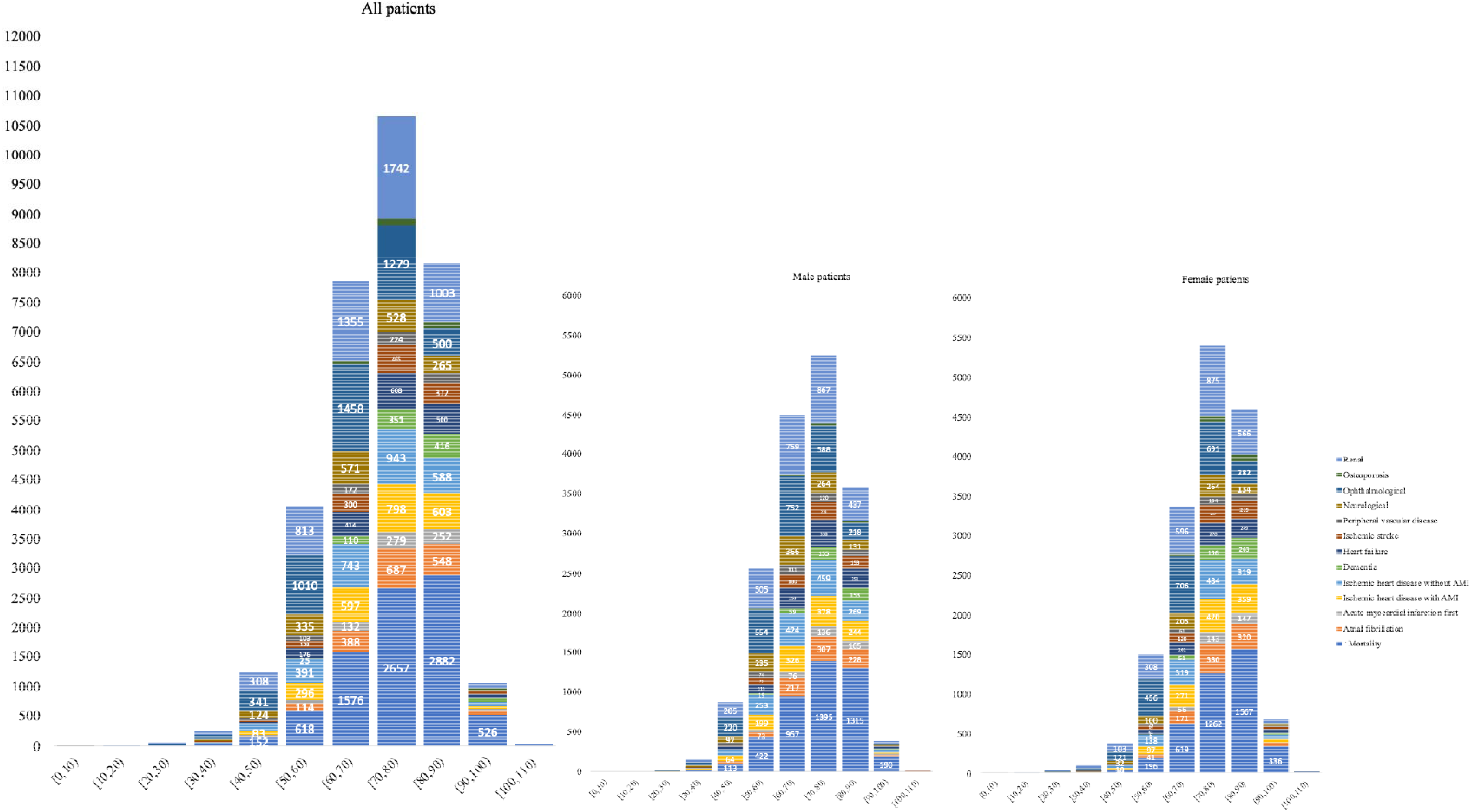
Statistics of complication onsets for all, male, and female groups at different age intervals.

### Significant complications to predict mortality

In univariable analysis with mortality as prediction outcome (**Table 2**), the odds of heart failure (odds ratio: 16.1, 95% CI: [12.9,19.9]) and AMI without known IHD (odds ratio: 2.5, 95% CI: [2.1,3.0]) were higher than those for other complications: ischemic heart disease with AMI (odds ratio: 2.3, 95% CI: [2.1,2.6]), peripheral vascular disease (odds ratio: 2.3, 95% CI: [1.9,2.8]), atrial fibrillation (odds ratio: 1.7, 95% CI: [1.5,1.8]), dementia (odds ratio: 1.8, 95% CI: [1.6,2.1]), ischemic stroke (odds ratio: 1.4, 95% CI: [1.2,1.6]), renal complication (odds ratio: 1.4, 95% CI: [1.3,1.5]). Ischemic heart disease without AMI (odds ratio: 1.0, 95% CI: [0.9,1.0], p value: 0.3051), neurological complication (odds ratio: 0.9, 95% CI: [0.8,1.0], p value: 0.0082), osteoporosis complication (odds ratio: 0.9, 95% CI: [0.7,1.1], p value: 0.1932) were not significant to predict mortality. In addition, male gender (odds ratio: 1.1, 95% CI: [1.0,1.2], p value: 0.0018) is predictive.

**Table 2.** Univariable regression for mortality prediction.

\* for $p \leq 0.05$ , \*\* for $p \leq 0.01$ , \*\*\* for $p \leq 0.001$
| Variable | Odds ratio (95% CI) | P-value |
| --- | --- | --- |
| Gender (male) | 1.1[1.0,1.2] | 0.0018** |
| Atrial fibrillation | 1.7[1.5,1.8] | <0.0001*** |
| AMI without known IHD | 2.5[2.1,3.0] | <0.0001*** |
| IHD with AMI | 2.3[2.1,2.6] | <0.0001*** |
| IHD without AMI | 1.0[0.9,1.0] | 0.3051 |
| Dementia | 1.8[1.6,2.1] | <0.0001*** |
| Heart failure | 16.1[12.9,19.9] | <0.0001*** |
| Ischemic stroke | 1.4[1.2,1.6] | <0.0001*** |
| Peripheral vascular disease | 2.3[1.9,2.8] | <0.0001*** |
| Neurological | 0.9[0.8,1.0] | 0.0082 |
| Ophthalmological | 0.5[0.4,0.5] | <0.0001*** |
| Osteoporosis | 0.9[0.7,1.1] | 0.1932 |
| Renal | 1.4[1.3,1.5] | <0.0001*** |

Significant variables (p value<0.001) were then used as input of multivariate logistic regression. The results (**Table 3**) show that HF (odds ratio: 16.6, 95% CI: [13.0, 20.1]), AMI without known IHD (odds ratio: 2.8, 95% CI: [2.3, 3.4]), peripheral vascular disease (odds ratio: 2.3, 95% CI: [1.9, 2.8]), dementia (odds ratio: 2.1, 95% CI: [1.8, 2.4]), and ischemic heart disease with AMI (odds ratio: 2.4, 95% CI: [2.1, 2.6]) were the most important predictors of mortality outcome.

**Table 3.**
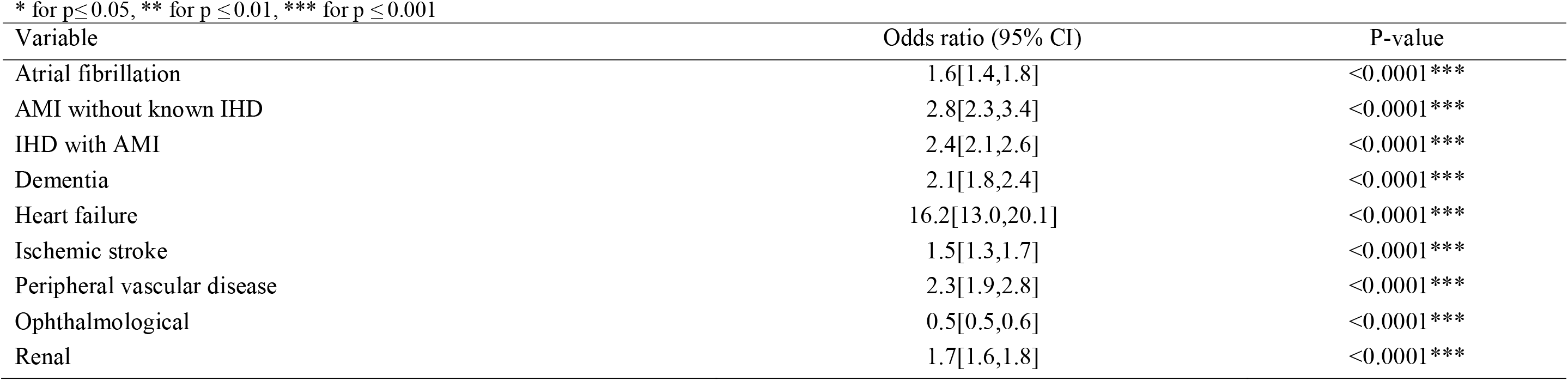
Multivariable regression for mortality prediction.

\* for $p \leq 0.05$ , \*\* for $p \leq 0.01$ , \*\*\* for $p \leq 0.001$
| Variable | Odds ratio (95% CI) | P-value |
| --- | --- | --- |
| Atrial fibrillation | 1.6[1.4,1.8] | <0.0001*** |
| AMI without known IHD | 2.8[2.3,3.4] | <0.0001*** |
| IHD with AMI | 2.4[2.1,2.6] | <0.0001*** |
| Dementia | 2.1[1.8,2.4] | <0.0001*** |
| Heart failure | 16.2[13.0,20.1] | <0.0001*** |
| Ischemic stroke | 1.5[1.3,1.7] | <0.0001*** |
| Peripheral vascular disease | 2.3[1.9,2.8] | <0.0001*** |
| Ophthalmological | 0.5[0.5,0.6] | <0.0001*** |
| Renal | 1.7[1.6,1.8] | <0.0001*** |

### Sequential complication patterns

We provide an illustrative explanation about the basic concept of the proposed KCPT+ in **Figure 2**, in which the sequence weighting scheme aims to discriminate the onset probability of the common and uncommon complication sequence. In this way, the model can accurately predict the next outcome of a given sequence in a scalable way by considering prior knowledge about sequence onset probability and preserving the advantage of CPT+’s lossless property to capture subsequence similarities.

**Figure 2.**
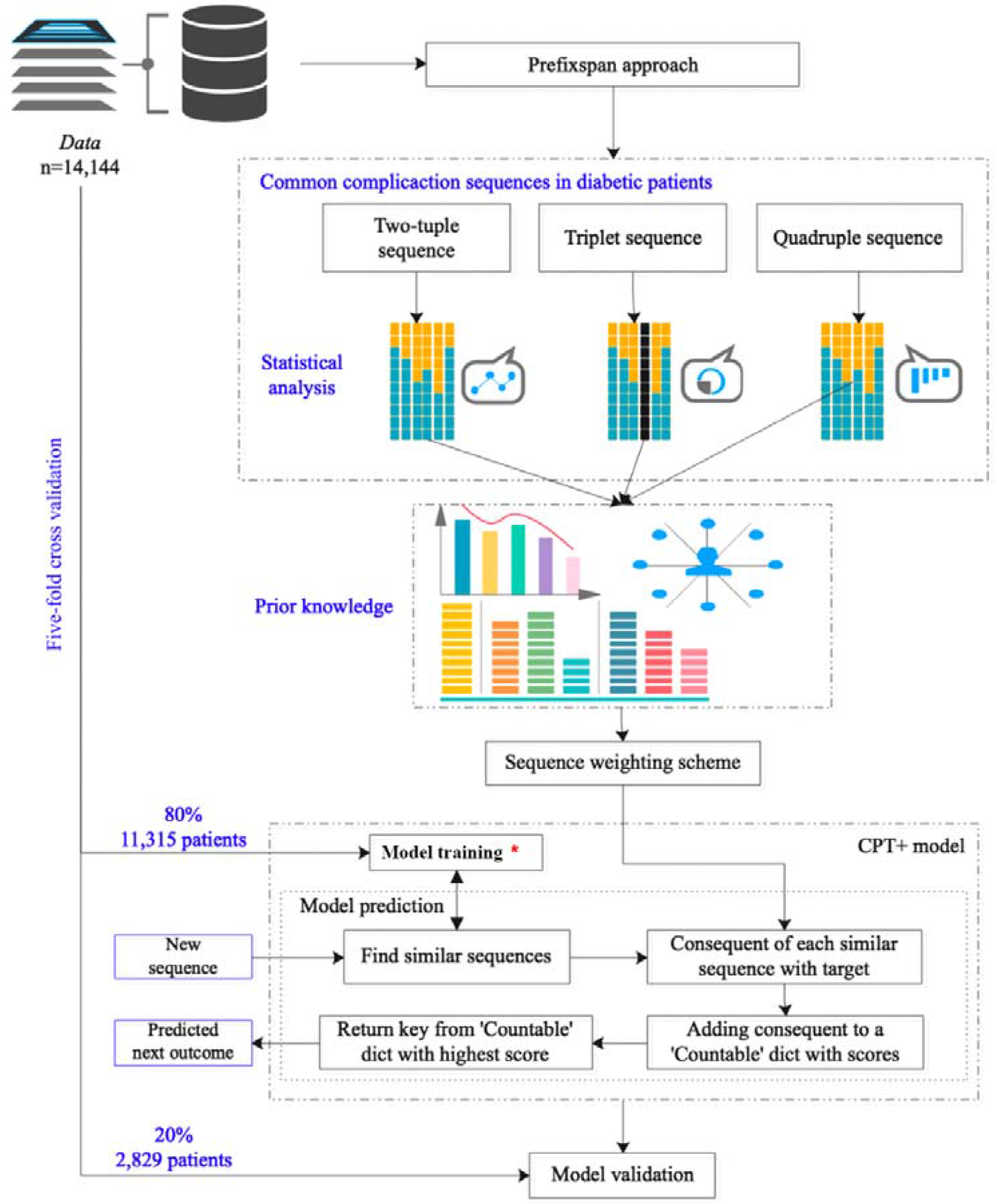
Basic concept of the proposed KCPT+ sequence prediction model. *: construct prediction tree, invested index, and lookup table from dataset.

We extract the sequential complication patterns of the 14, 144 patients with Prefixspan approach which identifies the couple, triplet, quadruple, quintuple, and sextuple sequential patterns according to the onset age of complications. The trajectories of complications are shown in **Figure 3**, which provides an easy-for-understanding graphical representation of the sequential complication patterns in diabetes mellitus. A wider line indicates more patients experienced that directed pairwise complication sequence with total patient number marked on the corresponding sequence edges. A Sankey diagram visualizes the proportional flow between complications within the pathology network. The Sankey network is used to illustrate the pathology development of diabetes mellitus complication patterns (**Figure 4**) with a corresponding number of patients who experienced that complication development (wider grey lines indicates more patients).

**Figure 3.**
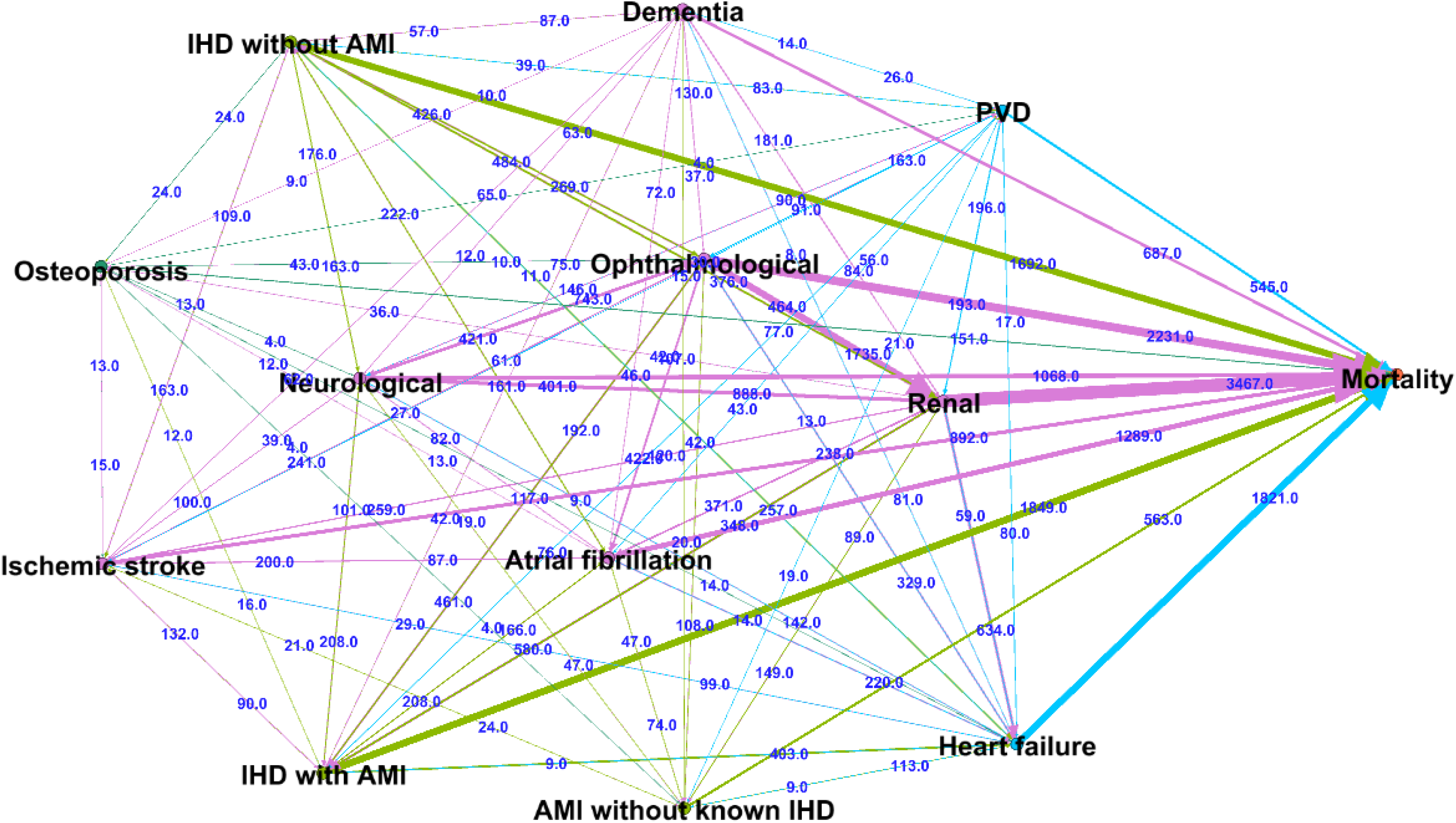
Graphical representation of sequential complication patterns in diabetes mellitus.

**Figure 4.**
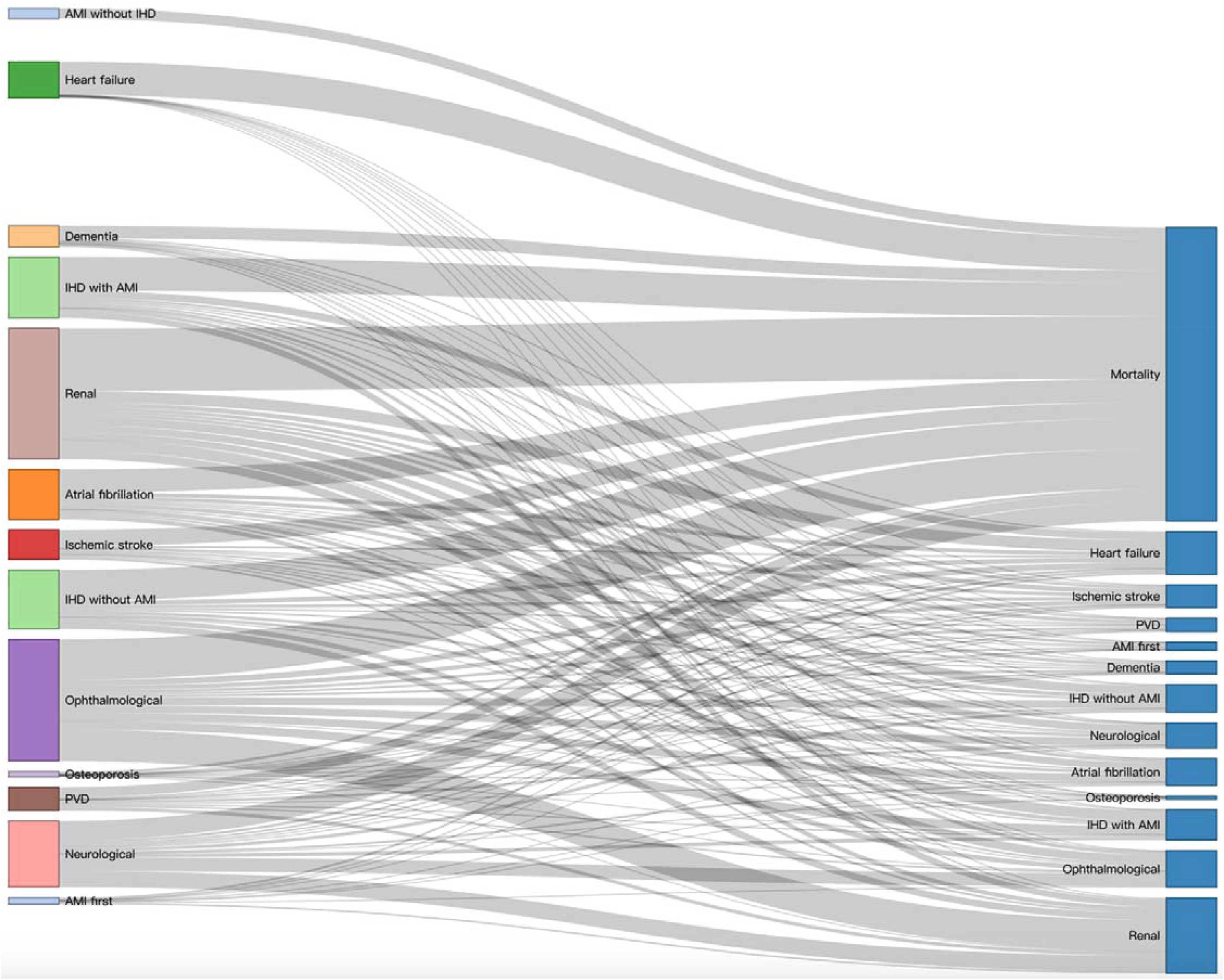
Sankey network to illustrate the pathology development of diabetes mellitus complication patterns.

#### (1) Couple sequences

The top 20 most frequent couple sequences are shown in **Table 4**. A total of 8491 patients died during the study period, and all had at least one complication. Among the couple sequences with mortality as the destination, renal complication was the commonest (n=3467), followed by ophthalmological complication (n=2231), ischemic heart disease with AMI (n=1849), heart failure (n=1821), ischemic heart disease without AMI (n=1692), atrial fibrillation (n=1289), neurological complications (n=1068), ischemic stroke (n=892), AMI without known IHD (n=563) and peripheral vascular disease (n=545).

**Table 4.**
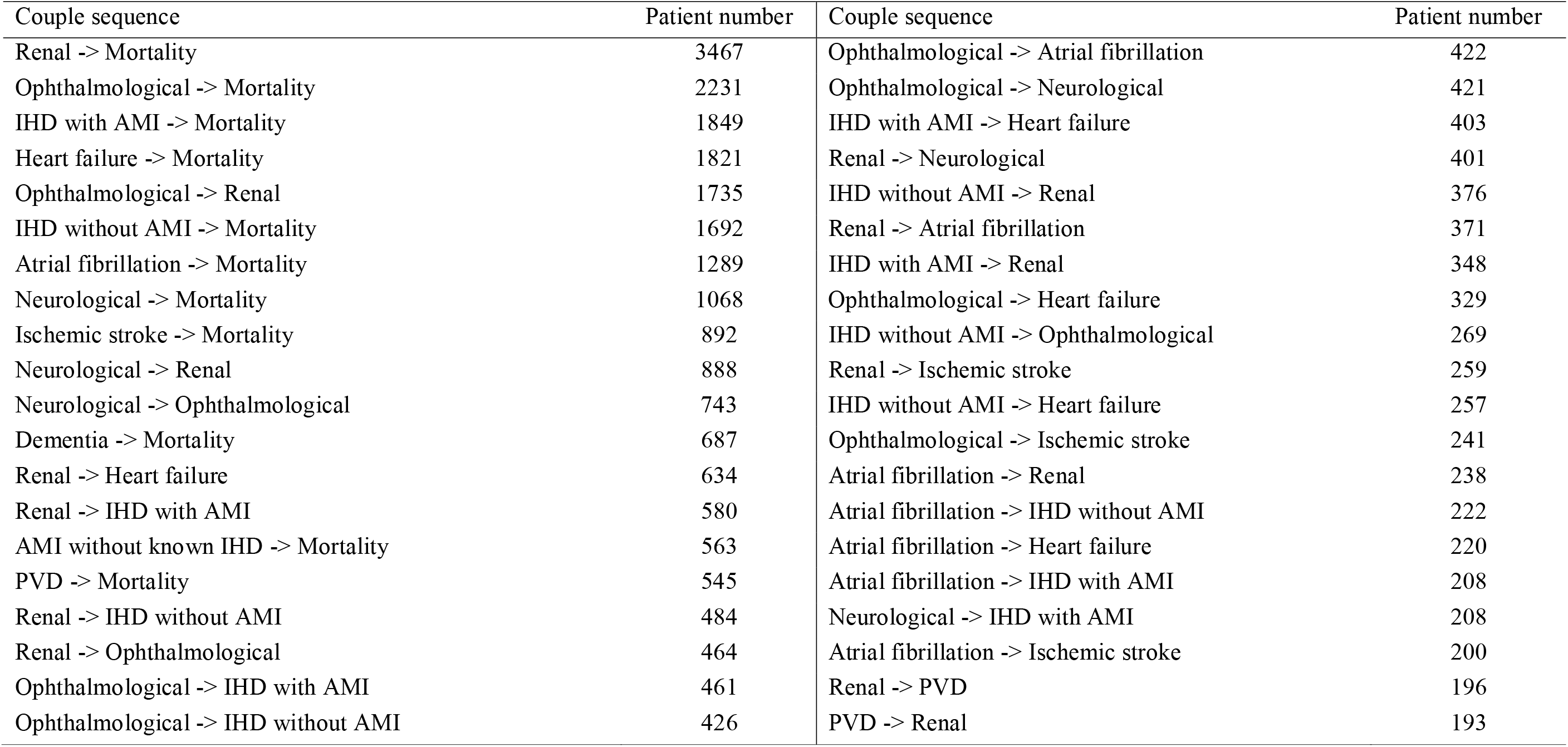
Couple sequence patterns (top 20)

The commonest complication couple sequences were: ophthalmological complications → renal complications (n=1735), neurological complications → renal complications (n= 888), neurological complications → ophthalmological complications (n=743), renal complications → heart failure (n=643), renal complications → IHD with AMI (n=580), renal complications → IHD without AMI (n=484), renal complications → ophthalmological complications (n=464), ophthalmological complications → IHD with AMI (n=461) and ophthalmological complications → IHD without AMI (n=426). Note that patients may have multiple complications at the same age.

#### (2) Triple sequences

Neurological → ischemic stroke → dementia is the most common triple sequence (n=930) in the cohort (**Table 5**), followed by: neurological → ophthalmological → ischemic stroke (n=493), renal → heart failure → mortality (n=462), IHD with AMI → osteoporosis → ischemic stroke (n=447), neurological → ophthalmological → renal (n=399), Renal → Neurological → Ischemic stroke (n=349).

**Table 5.**
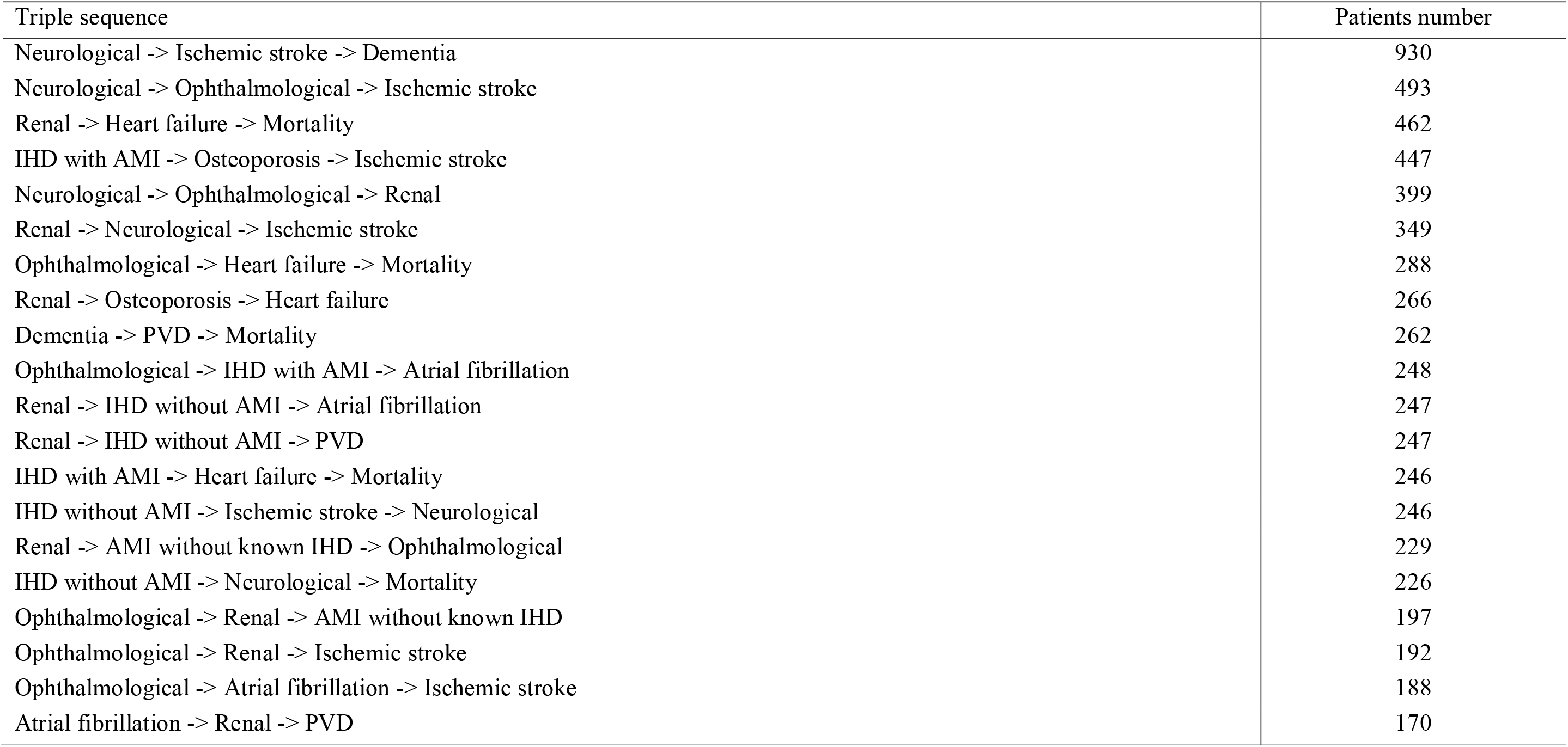
Triple sequence patterns (top 20)

#### (3) Quadruple sequences

We also identified quadruple complication sequence patterns of the diabetes mellitus patients as shown in **Table 6**. The most frequent sequence was dementia → IHD with AMI → heart failure → mortality (n=243), followed by ophthalmological → renal → heart failure → mortality (n=131), neurological → ophthalmological → renal → mortality (n=119), ophthalmological → renal → IHD with AMI → mortality (n=100), renal → IHD with AMI → heart failure → mortality (n=87).

**Table 6.** Quadruple sequence patterns (top 20)

| Quadruple sequence | Patients number |
| --- | --- |
| Dementia -> IHD with AMI -> Heart failure -> Mortality | 243 |
| Ophthalmological -> Renal -> Heart failure -> Mortality | 131 |
| Neurological -> Ophthalmological -> Renal -> Mortality | 119 |
| Ophthalmological -> Renal -> IHD with AMI -> Mortality | 100 |
| Renal -> IHD with AMI -> Heart failure -> Mortality | 87 |
| Ophthalmological -> Renal -> Atrial fibrillation -> Mortality | 81 |
| Neurological -> Renal -> IHD with AMI -> Mortality | 80 |
| Ophthalmological -> Renal -> Neurological -> Mortality | 75 |
| Ophthalmological -> Renal -> IHD without AMI -> Mortality | 73 |
| Neurological -> Renal -> Heart failure -> Mortality | 69 |
| Ophthalmological -> IHD with AMI -> Heart failure -> Mortality | 60 |
| Ophthalmological -> Renal -> Ischemic stroke -> Mortality | 58 |
| IHD with AMI -> Ophthalmological -> Renal -> Mortality | 56 |
| IHD without AMI -> Ophthalmological -> Renal -> Mortality | 55 |
| Neurological -> Ophthalmological -> IHD with AMI -> Mortality | 53 |
| Neurological -> Renal -> IHD without AMI -> Mortality | 53 |
| Neurological -> Ophthalmological -> Renal -> IHD without AMI | 50 |
| Ophthalmological -> Neurological -> Renal -> Mortality | 50 |
| Renal -> IHD without AMI -> Heart failure -> Mortality | 50 |
| Neurological -> Ophthalmological -> IHD without AMI -> Mortality | 49 |

#### (4) Quintuple and sextuple sequences

The identified most common ten quintuple and sextuple sequence patterns are included in **Tables 7** and **Table 8**, respectively. The commonest quintuple complication sequence was ischemic stroke → dementia → IHD without AMI → heart failure → mortality (n=28), followed by neurological → ophthalmological → renal → IHD without AMI → mortality (n=28) and ophthalmological → renal → IHD with AMI → heart failure → mortality (n=26).

For sextuple sequences, the commonest was neurological → ophthalmological → renal → IHD with AMI → heart failure → mortality (n=5). More detailed results of the complete couple, triple, quadruple, quintuple and sextuple sequence patterns are provided in **Supplementary Materials**. Patients with seven or more pathologies were not further analyzed, given the small numbers observed.

**Table 7.**
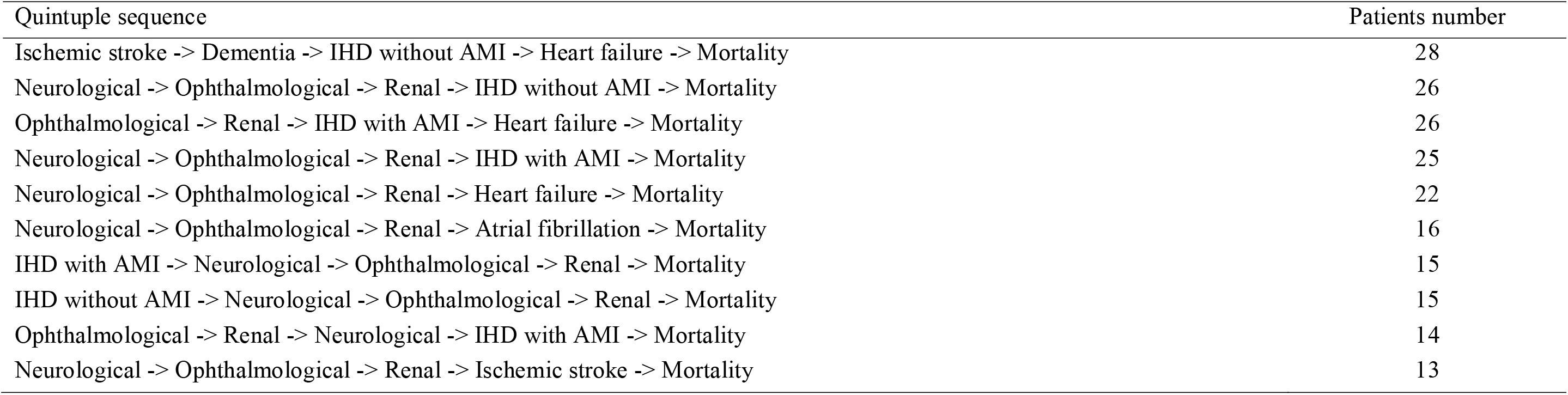
Quintuple sequence patterns (top 10)

**Table 8.**
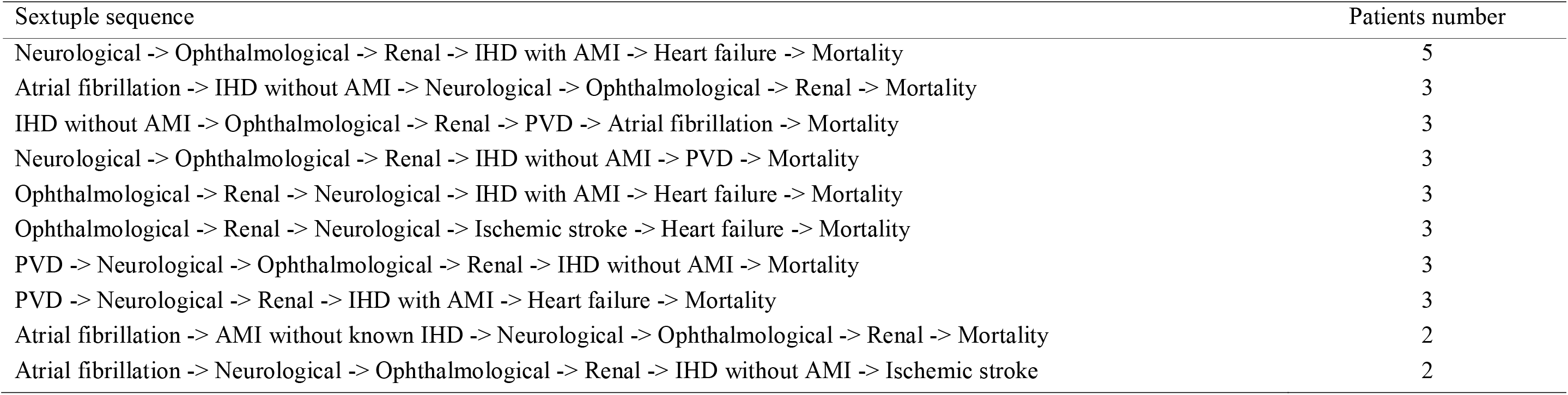
Sextuple sequence patterns (top 10)

| Sextuple sequence | Patients number |
| --- | --- |
| Neurological -> Ophthalmological -> Renal -> IHD with AMI -> Heart failure -> Mortality | 5 |
| Atrial fibrillation -> IHD without AMI -> Neurological -> Ophthalmological -> Renal -> Mortality | 3 |
| IHD without AMI -> Ophthalmological -> Renal -> PVD -> Atrial fibrillation -> Mortality | 3 |
| Neurological -> Ophthalmological -> Renal -> IHD without AMI -> PVD -> Mortality | 3 |
| Ophthalmological -> Renal -> Neurological -> IHD with AMI -> Heart failure -> Mortality | 3 |
| Ophthalmological -> Renal -> Neurological -> Ischemic stroke -> Heart failure -> Mortality | 3 |
| PVD -> Neurological -> Ophthalmological -> Renal -> IHD without AMI -> Mortality | 3 |
| PVD -> Neurological -> Renal -> IHD with AMI -> Heart failure -> Mortality | 3 |
| Atrial fibrillation -> AMI without known IHD -> Neurological -> Ophthalmological -> Renal -> Mortality | 2 |
| Atrial fibrillation -> Neurological -> Ophthalmological -> Renal -> IHD without AMI -> Ischemic stroke | 2 |

### Properties of the disease complication network

We conduct disease complication network analysis and calculated statistical properties (**Table 9**). In terms of properties of degree connection in the directed pathology network, renal, ophthalmological complications, atrial fibrillation, neurological complications, ischemic stroke, dementia have the largest values of in-degree (all with 12) and out-degree (all with 23), followed by heart failure and peripheral vascular disease both with in-degree (11) and out-degree (22), implying their important ‘intermediate’ role in the network. However, mortality as the destination has the largest out-degree value (12). The average degree of the complication network is 10.462.

**Table 9.** Properties of the diabetes mellitus pathology network.

| Outcome | Degree (In, Out) | Clo C | Har C | ECC | Bet C | Eig C | Hub | PageRank | Clu co |
| --- | --- | --- | --- | --- | --- | --- | --- | --- | --- |
| Renal | 11(12, 23) | 1.00 | 1.00 | 1 | 0.89 | 0.91 | 0.30 | 0.08 | 0.86 |
| Mortality | 12(0, 12) | 0.00 | 0.00 | 0 | 0.00 | 1.00 | 0.00 | 0.09 | 0.94 |
| Ophthalmological | 11(12, 23) | 1.00 | 1.00 | 1 | 0.89 | 0.91 | 0.30 | 0.08 | 0.86 |
| IHD with AMI | 9(10, 19) | 0.86 | 0.92 | 2 | 0.22 | 0.78 | 0.26 | 0.07 | 0.88 |
| Heart failure | 11(11, 22) | 0.92 | 0.96 | 2 | 0.67 | 0.91 | 0.28 | 0.08 | 0.86 |
| IHD without AMI | 9(10, 19) | 0.86 | 0.92 | 2 | 0.22 | 0.78 | 0.26 | 0.07 | 0.88 |
| Atrial fibrillation | 11(12, 23) | 1.00 | 1.00 | 1 | 0.89 | 0.91 | 0.30 | 0.08 | 0.86 |
| Neurological | 11(12, 23) | 1.00 | 1.00 | 1 | 0.89 | 0.91 | 0.30 | 0.08 | 0.86 |
| Ischemic stroke | 11(12, 23) | 1.00 | 1.00 | 1 | 0.89 | 0.91 | 0.30 | 0.08 | 0.86 |
| Dementia | 11(12, 23) | 1.00 | 1.00 | 1 | 0.89 | 0.91 | 0.30 | 0.08 | 0.86 |
| AMI without known IHD | 9(10, 19) | 0.86 | 0.92 | 2 | 0.22 | 0.78 | 0.26 | 0.07 | 0.88 |
| PVD | 11(11, 22) | 0.92 | 0.96 | 2 | 0.67 | 0.91 | 0.28 | 0.08 | 0.86 |

Several centrality measures were calculated. Firstly, closeness centrality was the largest for renal and ophthalmological complications, AF, neurological complications, ischemic stroke, dementia (all equal to 1.00), followed by HF and peripheral vascular disease (all equal to 0.9), implying their closeness importance in the network. This can be further confirmed by the same results with harmonic closeness centrality which calculates almost the same results. Secondly, eccentricity centrality can be interpreted as the easiness of a complication to be reached by all other complications in the network. IHD with AMI, heart failure, IHD without AMI, AMI without known IHD, and peripheral vascular disease have the largest eccentricity value (all equal to 2), indicating these complications are more easily reachable in the pathology network. Thirdly, the betweenness centrality was largest for renal and ophthalmological complications, AF, neurological, ischemic stroke, and dementia (all equal to 0.89), followed by HF and peripheral vascular disease (both with 0.67), implying that they can easily reach others on relatively short paths and lie on considerable fractions of shortest paths connecting others. The ranking results of eigen centrality calculations are almost the same with the betweenness centrality measure, except that ophthalmological also ranks the highest with eigen centrality value as 0.91.

Finally, PageRank value was the highest for mortality (0.09), followed by renal and ophthalmological complications, HF, AF, neurological, ischemic stroke, dementia, and peripheral vascular disease (all equal to 0.08). The clustering coefficient can be used to detect whether complications tend to create tightly knit groups characterized by a relatively high density of connections. Mortality has the largest clustering coefficient (0.94), followed by IHD with or without AMI, and AMI without known IHD (all equal to 0.88). This implies that they tend to form a clique with other neighbor complications in the pathology of diabetes mellitus. The average clustering coefficient of the comorbidity pathology network is 0.87.

The identified sequential patterns provide evidence for identifying diabetes mellitus complication development and shows promising clinical and medical value for diabetes mellitus treatment optimization and even reduce overall mortality.

### Sequence prediction results

The proposed KCPT+ sequence prediction model was employed to predict the next possible outcome of patients with diabetes mellitus. The dataset with 14,144 patients is randomly split in a five-fold cross-validation way into training dataset (80%, 11,315 patients) and validation dataset (20%, 2,829 patients). We trained all sequence prediction models and then compare their prediction performance on the validation dataset (**Table 10**). The proposed KCPT+ model outperforms the CPT+ model and other baselines including RNN and LSTM according to evaluation metrics, implying that consideration of prior knowledge about the probabilities of sequence patterns can significantly improve model’s overall sequence prediction ability.

**Table 10.**
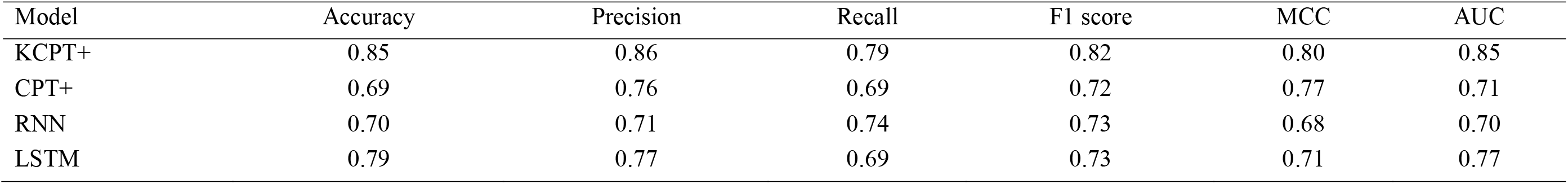
Comparative performance evaluation of sequence prediction models on validation dataset (all sequence). AUC: area under the curve. MCC: Matthews correlation coefficient.

Besides, we perform KCPT+ model on separate sequence outcome datasets to predict the primary outcomes with previous complication sequences as input. The results (**Table 11**) show that the model gains the best performance to predict mortality (F1 score 0.90, ACU 0.88), osteoporosis (F1 score 0.86, AUC 0.82), ophthalmological complication (F1 score 0.82, AUC 0.82), IHD with AMI (F1 score 0.81, AUC 0.85), neurological complication (F1 score 0.81, AUC 0.83). The experiment results demonstrate that the proposed model can efficiently predict primary sequence outcomes of diabetes mellitus patients with high accuracy. The model shows the potential to early diagnosis of possible complications and mortality onset based on patients’ previous disease sequences as the core module of medical assistant decision systems for healthcare use.

**Table 11.** Model performance of KCPT+ in predicting primary outcomes. AUC: area under the curve. MCC: Matthews correlation coefficient.

| Sequence outcome | Accuracy | Precision | Recall | F1 score | MCC | AUC |
| --- | --- | --- | --- | --- | --- | --- |
| Mortality | 0.89 | 0.91 | 0.89 | 0.90 | 0.83 | 0.88 |
| Osteoporosis | 0.88 | 0.85 | 0.88 | 0.86 | 0.83 | 0.82 |
| Dementia | 0.81 | 0.70 | 0.80 | 0.74 | 0.78 | 0.73 |
| AMI without known IHD | 0.86 | 0.93 | 0.69 | 0.79 | 0.76 | 0.82 |
| Atrial fibrillation | 0.79 | 0.84 | 0.68 | 0.75 | 0.86 | 0.80 |
| Heart failure | 0.79 | 0.73 | 0.67 | 0.70 | 0.80 | 0.73 |
| Ischemic stroke | 0.84 | 0.81 | 0.79 | 0.80 | 0.75 | 0.74 |
| IHD with AMI | 0.75 | 0.85 | 0.78 | 0.81 | 0.85 | 0.85 |
| IHD without AMI | 0.85 | 0.81 | 0.71 | 0.76 | 0.71 | 0.78 |
| Peripheral vascular disease | 0.76 | 0.72 | 0.74 | 0.73 | 0.66 | 0.78 |
| Renal | 0.85 | 0.75 | 0.83 | 0.79 | 0.85 | 0.84 |
| Neurological | 0.84 | 0.79 | 0.84 | 0.81 | 0.78 | 0.83 |
| Ophthalmological | 0.73 | 0.86 | 0.79 | 0.82 | 0.90 | 0.82 |

## Discussion

In this study, we developed a knowledge enhanced CPT+ (KCPT+) model which considers previously known onset probability of couple, triple, and quadruple sequences to further improve overall prediction ability while at the same time preserve the advantage of CPT+ to capture the subsequence similarities without information loss. The main findings of this study are summarized as follows:

1. the median onset age in diabetic patients were identified: ophthalmological complication occurs at the earliest, followed by neurological and renal complications.
2. the commonest couple, triple, quadruple, quintuple, and sextuple sequence patterns in diabetic patients were identified. Easy-for-understanding graphical representation of the sequential complication patterns is presented to identify typical progression trajectories of diabetes-related complications.
3. network analyses were conducted to extract meaningful comorbidity connection properties, identifying meaningful clusters of comorbidities that tend to occur together.
4. an accurate sequence prediction model was developed for predicting the next possible complication (or mortality) with any given prior sequence. The proposed KCPT+ model outperforming other models including CPT, CNN, and LSTM. The sequence prediction model can help clinicians to devise effective treatment strategies for diabetes-related complications before they develop.

Sequential pattern analysis has been applied in order to aid decision making for changing the treatment dose of insulin in type 1 diabetics [28] and to predict the next prescribed medication for diabetes [29]. In terms of trajectory analysis of disease patterns in diabetes, the study from Korea demonstrated progression trajectory from 1) retinopathy → polyneuropathy → peripheral vascular disease, and 2) depressive episode → musculoskeletal disorders → thyroid disorders [9]. By contrast, the study from Denmark found a total of 1,171 significant trajectories. These authors grouped these into patterns centred on key diagnoses such as chronic obstructive pulmonary disorder and gout, which they found to be central for disease progression [10]. In our study, we focused on the trajectory pattern of specifically diabetes-related complications, revealing several important trajectory sequences up to six sequential complications.

Compact prediction tree plus (CPT+) [24] has been proposed as a fairly new probabilistic predictive model to assist sequential pattern analysis. The fundamental advantage of CPT+ is that it compresses training sequences without information loss by exploiting similarities between subsequences and is working with low time complexity. Existing studies have shown that CPT+ outperforms traditional sequence mining approaches including prediction by partial matching (PPM) [30], all-kth order Markov model (AKOM) [31], dependency graphs (DG) [32], transition directed acyclic graph (TDAG) [33]. Traditional sequence prediction models make the Markovian assumption that each event solely depends on previous events. This may lead to reduced prediction accuracy [34], i.e., these traditional models are built using only part of the information contained in training sequences (Markov models typically considers only the last k items of training sequences to perform a prediction, where k is the order of the model). However, increasing the order of Markov models often induces a very high state complexity, thus making the model impractical for real applications [35]. Consideration of complete information contained in training sequences (sequential patterns not just dependent on previous events) is expected to improve the overall sequence prediction performance. CPT+ considers the subsequence similarity information to improve prediction accuracy with low time complexity.

### Limitations

Several limitations of this study should be noted. Firstly, as this was an administrative database study, -coding and coding error is a possibility. Secondly, given the retrospective nature of this study, missing data may lead to information bias.

## Conclusion

This study provides analyses about the sequential pattern characteristics of disease-related complications that adversely affect human’s quality-of-life. The identified couple, triple, quadruple, quintuple, and sextuple sequence patterns benefit the understanding of the complication development pathology. The proposed accurate complication sequence prediction model can be implemented as a core module of a medical assistant decision system for better risk-stratified care, to enable early complication detection and prevention.

## Data Availability

The anonymized dataset has been deposited in the following repository's URL:
https://zenodo.org/record/4382440#.X-DRTNgzaUl

